# Multi-omics and Mendelian Randomization network analysis of the association between metabolic and cognitive functions in the UK Biobank Database

**DOI:** 10.1101/2024.05.30.24307732

**Authors:** Paolo Alberto Lorenzini, Sarah Raye Langley

## Abstract

In obese individuals, low levels of leptin, an hormone regulating energy expenditure, have been claimed to be associated with neurodegeneration during ageing. Since leptin signalling system is less potent in the elderly, its protective anti-inflammatory effect in the brain is inhibited and people overweight in middle-age are at higher risk of developing dementia later in life. However, the causal association between obesity and cognitive decline has been inconsistent so far. We incorporated several metabolic and cognitive phenotypes from primary-care health records from European individuals in the UK Biobank to study comprehensively the genetic architecture of metabolic function and cognition. By GWAS analysis, we identified genetic loci in various traits associated with metabolic function and cognitive performance and investigated their genetic association by Mendelian Randomization. Importantly, we identified putatively causality amongst metabolism related traits and that higher adiposity is causally associated with a worse cognitive performance. We conducted a transcriptome-wide association study to identify regulatory effects of the susceptibility variants to pinpoint genes associated with gene expression changes. We integrated the GWAS results with transcriptome and single cell public data to identify tissues and cell types associated with the pathogenesis of metabolic dysfunction and cognition. Overall, this *multi*-*omics* approach have proven to be invaluable for demonstrating the causal association between adiposity and cognition and for the discovery of causal variants, genes, tissues and cell types underlying these phenotypes.

## Introduction

Increases in life expectancy coupled with decreases in fertility rates worldwide are leading towards an aging society (WHO, 2020). As a person grows older, the cognitive functions can deteriorate leading to difficulties with a person’s processing, remembering and concentrating^1^. A certain baseline level of cognitive decline is normal during aging. However, when the cognitive impairment has become severe enough to compromise the ability to perform activities of daily life, dementia is diagnosed^2,3^. As the global population ages, more people will be affected by cognitive decline and more likely at risk to develop dementia^4,5^. According to recent reports, there are about 55 million people currently living with dementia and it is forecasted that cases will triple by 2050^6^. This projected trend underscores the need for public health planning efforts and public policies to promote healthy aging and reduce the burden that an aging population is posing on society^5–7^.

In addition to developing therapeutics, in order to develop pre-emptive strategies, it is very important to assess which are the modifiable risk factors that could potentially exacerbate, or reduce, one’s risk of developing dementia in later life^8^. Type 2 diabetes (T2D), hypertension, and obesity (i.e. metabolic traits) as well as depression, smoking and low educational attainment have been identified as exacerbating factors of dementia^9–11^. In particular, epidemiological studies have demonstrated that T2D can increase the risk of cognitive impairment and that patients with T2D are more likely to be diagnosed with dementia^12–14^. Furthermore, obesity, T2D and dementia share several biological features such as chronic inflammation, severe oxidative stress and impairment in insulin and energy metabolism^13,15–17^.

Given this relationship among obesity, T2D, and dementia, understanding the mechanistic links among them is essential to developing effective strategies for prevention and treatment of these diseases^18,19^. Despite this strong observational evidence that T2D is associated with dementia, causality cannot be reliably established since these observational studies are prone to confounding effects and reverse causation. Mendelian Randomization (MR) is an alternative approach to randomized clinical trials (RCTs) that can be utilised to determine causality by circumventing confounding effects and reverse causation^20,21^. MR is an epidemiological methodology which uses genetic variants, as instrumental variables (IVs), in order to determine causal relationship between a modifiable risk factor and an outcome^20,22,23^. Because of the random allocation of alleles at conception, genetic variants associated with a modifiable exposure are randomly distributed in relation to potential confounders. Furthermore, considering that the genotype inherited at birth stays constant throughout life, the possibility of reverse causation can be ruled out^20,22,23^. Importantly, in order to estimate direct causal effects, multivariable MR techniques can also be used to take into account of indirect effects due to “pleiotropy”, a widespread phenomenon in the genome by which a single SNP influences multiple traits^24^.

Previous MR studies have assessed the causality between T2D, insulin sensitivity and fasting glucose on the risk of neurological diseases with contradictory results. Several studies did not find significant associations between metabolic factors and dementias^25–27^, while others identified significant associations between glycaemic traits and increased risk of dementia and/or cognitive decline^28–31^. We conducted a GWAS analysis of traits related to metabolic function and cognitive performance in European individuals in the UK Biobank database and utilised the results to determine causality (Figure 1A-B).

**Figure 1.**
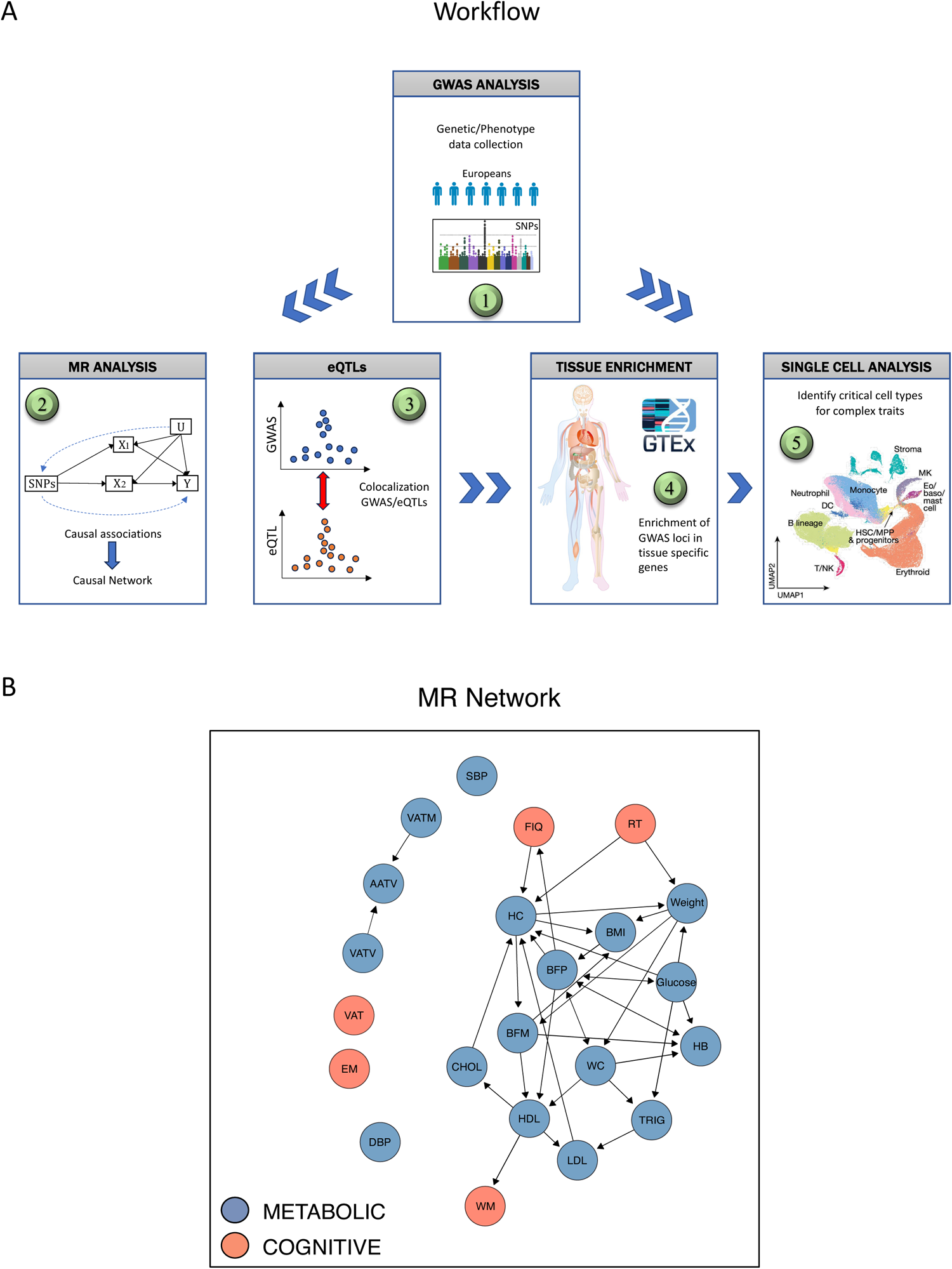
**(A)** Summary of the workflow conducted in this study. (1) GWAS analysis was performed on genetic and phenotypic data of individuals with European ancestry obtained from UK Biobank database. (2) The GWAS results were then utilized to determine causality by Mendelian Randomization (MR). Subsequently, GWAS were integrated with eQTLs gene expression data in skeletal muscle of individual of European ancestry to perform pleiotropy analysis (3). Finally, GWAS were integrated with bulk RNA-seq public data to perform tissue enrichment analysis (4) and with single cell RNA-seq public data to perform cell-type specific analysis (5). **(B)** Mendelian Randomization Network (MRN) of metabolic (blue) and cognitive (red) traits in Europeans. Single-headed arrows indicate direct causal effects. Bi-directional double-headed arrows indicate that traits causally influence each other. Systolic Blood Pressure (SBP), Diastolic Blood Pressure (DBP), Low Density Lipoproteins (LDL), High Density Lipoproteins (HDL), Waist Circumference (WC), Hip Circumference (HC), Cholesterol (CHOL), Triglycerides (TRIG), Body Mass Index (BMI), Glycated Hemoglobin (HB), Fat Mass (FM), Total Fat (TF), Total Fat Percentage (TFP), Visceral Adipose Tissue Volume (VATV), Visceral Adipose Tissue Mass (VATM), Abdominal Adipose Tissue Volume (AATV), Body Fat Mass (BFM), Body Fat Percentage (BFP), Working Memory (WM), Episodic Memory (EM), Fluid IQ (FIQ), Reaction Test (RT), Visual Attention Test (VAT). GWAS genome-wide association study, SMR summary data-based Mendelian randomization, eQTL expression quantitative trait loci.

While these genome-wide associations utilised in MR analyses provide a useful framework for identifying putative susceptibility loci, they rarely identify causal genes, predominantly due to the complicated Linkage Disequilibrium (LD) structure of the genome, as well as to the fact that genetic variants can affect phenotype via distant regulation of gene expression^32^. In order to prioritise functionally relevant genes from Genome-Wide Association Study (GWAS) loci for follow-up studies, we integrated the GWAS results with expression Quantitative Trait Loci (eQTLs) from human skeletal muscle tissue (Figure 1A), which allowed us to discover genetic variants associated with transcriptional changes^33^. In addition, we utilised this multi-omics analysis to assess for pleiotropy. We performed functional mapping and annotation to further explore genetic variants and genomic loci in the pathogenesis of metabolic dysfunction and cognitive decline.

Furthermore, the underlying cell types mediating predisposition to metabolic dysfunction remain largely obscure, despite identifying such cell types is a very important step towards a better understanding of mechanisms causing the disease^34^. For example, it has been recently reported that a dysregulation in specific adaptive immune cell types triggers the onset, development and progression of obesity and T2D^35^. As such, we integrated the GWAS results with public gene expression data in order to identify genes and cell types underlying susceptibility to obesity and cognitive health (Figure 1A). Overall, our multi-omics and MR analysis (Figure 1A) would provide important leads to better understand the pathogenesis of metabolic dysfunction and cognitive decline and to develop novel therapeutic targets for precision medicine.

## Results

### Establishing causal association between obesity and cognitive functions

We used genetic and phenotypic data from the UK Biobank Database (European cohort)^36^ to estimate the causal association between obesity and cognitive performance (Figure 1A-B). We selected several metabolic and cognitive traits (Table S1) to cover as many as possible phenotypes. To have a comprehensive assessment of cognitive health status, we selected cognitive traits covering all the four major cognitive domains (episodic and working memory, executive function, processing speed and attention)^37^ (Table S1). We obtained a final list of 22 traits (17 metabolic and 5 cognitive) (Table S1). For each trait, we performed a GWAS analysis, using both the entire population (sex-combined) or as individual sexes. Functional analysis of the sex-combined GWAS results confirmed that the significant hits (p-value < 5e-08) are involved in lipid metabolism and homeostasis (Supp. Fig. S1A). With the sex-split summary statistics, we built a Mendelian Randomization network (Figure 1B) by utilising an algorithm which allows to estimate direct causal effects not mediated by other measured factors^38^. Several associations were in line with previous epidemiological observations, with a few exceptions. Most of the direct causal associations (36/39) excluding some involving cholesterol and hip circumference showed an expected epidemiological association (Table S2). For example, an increase in waist circumference was causally associated with an increase in body fat percentage, glycated haemoglobin, hip circumference, triglycerides and a decrease in HDL (Figure 1B). Interestingly, an increase in body fat percentage was causally associated with a lower performance in the FIQ test (Figure 1B), assessing verbal and numerical reasoning (executive function domain). An increase in HDL was causally associated with a decrease in LDL and a higher number of digits remembered correctly in the working memory test and an higher time to correctly identifying matches in the reaction test assessing processing speed, was causally associated with an increase in waist circumference and weight (Figure 1B).

Collectively, these results show a strong causal association amongst metabolic traits related to adiposity and that metabolic function, in particular adiposity, is causally associated with a worse performance in certain cognitive test (Figure 1B, Table S2).

### Identifying pleiotropically causal genes in obesity and cognitive functions

In order to identify and prioritize functionally relevant causal genes in obesity or cognitive functions, we integrated the genetic data with gene expression data (cis-eQTLs) and conducted a pleiotropic gene-trait association analysis by SMR-HEIDI^39^. In brief, by combining cis-eQTL data from a specific tissue with GWAS summary statistics of a single trait, the SMR test allows to assess whether the effect size of a SNP on that specific trait is mediated by gene expression^39,40^. Importantly, the test allows as well to discriminate if the observed association could be due to a single variant or alternatively two distinct genetic variants in high LD with each other (pleiotropy)^33,39^.

We integrated the sex-combined GWAS summary statistics of the metabolic and cognitive traits with cis-eQTLs data from skeletal human muscle tissue obtained from the GTEx database^41^. We focused on skeletal muscle tissue in particular because obesity-linked insulin resistance is mainly due to fatty acid overload in non-adipose tissues, particularly skeletal muscle and liver^42^. Since 10282 eQTLs probes (1.000.905 SNPs) were included, the genome-wide significance level (P_SMR_) is defined as 0.05/10282 = 5.0 × 10^−6^.

We ran the SMR analysis with default settings (see methods). For the metabolic traits, we identified 140 significant probes tagging 82 unique genes which passed the P_SMR_ threshold, whereas for the cognitive traits only 4 genes/probes showed significant association (Tables S3-4).

Since significant SMR results could also reflect linkage (i.e. the top associated cis-eQTL being in LD with two distinct causal variants, one affecting gene expression and the other affecting trait variation), for the significant probes with P_SMR_ < 5.0 × 10^−6^, we conducted a HEIDI test. If a probe passes the HEIDI test (P_HEIDI_ ≥ 0.05), it signifies that there is a single causal variant affecting trait and gene expression (pleiotropy or causality), which is of more important biological interest and should be prioritized for follow-up studies. After application of the HEIDI test (P_HEIDI_ ≥ 0.05), we retained 62 probes (60 metabolic, 2 cognitive) tagging to 39 unique genes (37 metabolic, 2 cognitive) (Table S5). Functional analysis of these genes showed enrichment in lipid metabolism (Supp. Fig. S1B) and a Protein-Protein Interaction (PPI) network of these 39 genes showed strong evidence of interaction between some of those proteins, including those involved in lipid metabolism (Supp. Fig. S2A-B).

Interestingly, several of the significant probes which passed the P_SMR_ threshold (33/144), tagging 13 unique genes were located on a chromosome 16 region well known be associated in the pathogenesis of human obesity and neural development^43–46^ (Figure 2A-B, Table 1, Table S4).

**Figure 2.**
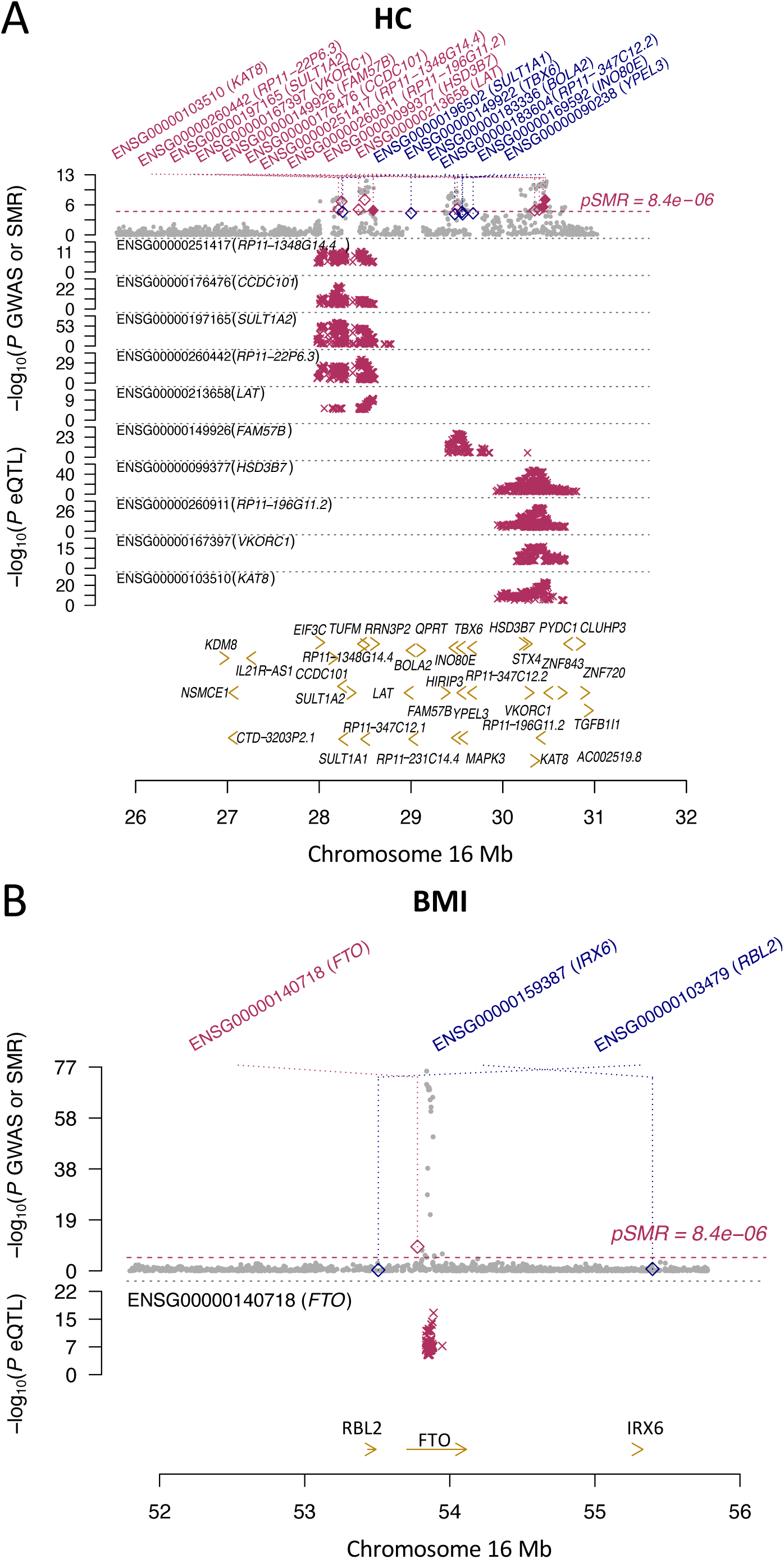
**(A)** SMR plot of the K(lysine) acetyltransferase 8 (KAT8) locus for the Hip Circumference (HC) GWAS trait in Europeans. **(B)** SMR plot of the Fat mass and obesity-associated (FTO) locus for the Body Mass Index (BMI) GWAS trait in Europeans. On the y-axis at the top, -log10(P values) for SNPs reported in this GWAS meta-analysis are represented by grey dots and rhombuses represent the P-values for probes from the reverse SMR test. Solid rhombuses are indicating that the probes passed the HEIDI test and hollow rhombuses indicating that the probes did not pass the HEIDI test. The red dashed line indicates significant pSMR threshold in the SMR test. In the middle plot, magenta x-crosses represent the cis-eQTL -log10(P values) of SNPs associated with gene expression in skeletal muscle (GTEx). Bottom plot, yellow arrows indicate genes over the genome positions in hg38 (x-axis). GWAS genome-wide association study, SMR summary data-based Mendelian randomization, HEIDI heterogeneity in dependent instruments, eQTL expression quantitative trait loci, GTEx Genotype-Tissue Expression.

**Table 1:** Most significant SMR probes in Europeans located on chromosome 16.

| Trait | Probe ID | Gene | Chr | Top SNP | Allele Freq | $P_{\text{eQTL}}$ | $P_{\text{GWAS}}$ | $P_{\text{SMR}}$ |
| --- | --- | --- | --- | --- | --- | --- | --- | --- |
| BMI | ENSG00000140718 | FTO | 16 | rs1477196 | 0.620664 | 2.95E-12 | 1.71E-39 | 7.03E-10 |
| CHOL | ENSG00000040199 | PHLPP2 | 16 | rs1058750 | 0.832374 | 6.47E-44 | 1.74E-10 | 6.62E-09 |
| HC | ENSG00000103510 | KAT8 | 16 | rs9925964 | 0.361484 | 3.03E-23 | 1.45E-11 | 2.35E-08 |
| HC | ENSG00000260442 | RP11-22P6.3 | 16 | rs8049439 | 0.405005 | 4.53E-35 | 4.11E-10 | 2.45E-08 |
| HC | ENSG00000197165 | SULT1A2 | 16 | rs4788084 | 0.419946 | 1.27E-61 | 9.81E-09 | 6.01E-08 |
| BFM | ENSG00000158865 | SLC5A11 | 16 | rs2287783 | 0.195463 | 9.03E-76 | 1.46E-07 | 4.28E-07 |
| HC | ENSG00000167397 | VKORC1 | 16 | rs2884737 | 0.258031 | 2.13E-16 | 9.17E-10 | 9.14E-07 |
| Weight | ENSG00000149926 | FAM57B | 16 | rs9928448 | 0.466783 | 2.51E-27 | 5.53E-08 | 1.20E-06 |
| BMI | ENSG00000103550 | KNOP1 | 16 | rs62025019 | 0.171962 | 3.17E-94 | 8.49E-07 | 1.68E-06 |
| Weight | ENSG00000176476 | CCDC101 | 16 | rs480400 | 0.543293 | 2.12E-25 | 8.00E-08 | 1.83E-06 |
| HDL | ENSG00000213398 | LCAT | 16 | rs1134760 | 0.163383 | 1.62E-11 | 2.09E-11 | 2.03E-06 |
| HC | ENSG00000251417 | RP11-1348G14.4 | 16 | rs8049439 | 0.405005 | 3.16E-12 | 4.11E-10 | 3.27E-06 |
| HC | ENSG00000260911 | RP11-196G11.2 | 16 | rs35468353 | 0.376089 | 4.28E-31 | 4.05E-07 | 3.44E-06 |

BMI Body Mass Index, CHOL Cholesterol, HC Hip Circumference, BFM, Body Fat Mass, HDL High Density Lipoproteins, Chr Chromosome number, Allele Freq Allele Frequency, $P_{\text{eQTL}}$ p-value of the top associated cis-eQTL of the probe, $P_{\text{GWAS}}$ GWAS p-value of the top cis-eQTL, $P_{\text{SMR}}$ p-value for gene-trait association from the SMR test, $P_{\text{HEIDI}}$ p-value from HEIDI test to indicate whether the gene-trait association is due to a single shared genetic variant (the smaller $P_{\text{HEIDI}}$ the more likely that there are more than one genetic variant).

In addition, for those 144 probes, we detected high SMR signals as well on chromosomes 1,2,6,11,19,20, which suggests an involvement of these chromosomes in the pathogenesis of human metabolic dysfunctions and neurological diseases as well (Table 2, Table S4).

**Table 2:** Top most significant SMR probes in Europeans located on chromosomes 1,2,6,11,19,20.

| Trait | Probe ID | Gene | Chr | Top SNP | Allele Freq | $P_{eQTL}$ | $P_{GWAS}$ | $P_{SMR}$ |
| --- | --- | --- | --- | --- | --- | --- | --- | --- |
| LDL | ENSG00000143126 | CELSR2 | 1 | rs7528419 | 0.22256 | 2.23E-76 | 1.12E-121 | 8.55E-48 |
| TRIG | ENSG00000115234 | SNX17 | 2 | rs715325 | 0.392272 | 7.11E-87 | 6.55E-26 | 1.55E-20 |
| TRIG | ENSG00000198522 | GPN1 | 2 | rs3749147 | 0.252445 | 6.86E-18 | 1.17E-53 | 5.37E-14 |
| LDL | ENSG00000110080 | ST3GAL4 | 11 | rs11220462 | 0.128581 | 2.38E-47 | 1.15E-16 | 6.48E-13 |
| LDL | ENSG00000134222 | PSRC1 | 1 | rs646776 | 0.777289 | 4.83E-14 | 1.23E-122 | 7.08E-13 |
| TRIG | ENSG00000149485 | FADS1 | 11 | rs174549 | 0.308738 | 2.79E-19 | 5.26E-31 | 1.30E-12 |
| HDL | ENSG00000172247 | C1QTNF4 | 11 | rs17788930 | 0.348354 | 1.36E-41 | 1.82E-16 | 2.05E-12 |
| HDL | ENSG00000100979 | PLTP | 20 | rs6065904 | 0.211698 | 4.45E-22 | 1.31E-24 | 2.11E-12 |
| HDL | ENSG00000109919 | MTCH2 | 11 | rs4752856 | 0.35156 | 1.40E-33 | 5.77E-17 | 6.02E-12 |
| LDL | ENSG00000175164 | ABO | 9 | rs8176719 | 0.346567 | 1.70E-86 | 3.15E-13 | 8.16E-12 |
| LDL | ENSG00000142252 | GEMIN7 | 19 | rs3178166 | 0.494128 | 3.74E-52 | 4.21E-14 | 1.34E-11 |
| HB | ENSG00000179344 | HLA-DQB1 | 6 | rs3828790 | 0.609058 | 0 | 1.06E-11 | 2.05E-11 |
| HC | ENSG00000065060 | UHRF1BP1 | 6 | rs9462026 | 0.327944 | 4.31E-79 | 3.64E-12 | 7.01E-11 |
| HB | ENSG00000232629 | HLA-DQB2 | 6 | rs3828790 | 0.609058 | 3.19E-117 | 1.06E-11 | 7.06E-11 |
| TRIG | ENSG00000134824 | FADS2 | 11 | rs174538 | 0.315157 | 7.00E-15 | 2.13E-29 | 1.53E-10 |
| HDL | ENSG00000134575 | ACP2 | 11 | rs2167079 | 0.298227 | 1.00E-20 | 5.44E-17 | 4.52E-10 |
| Weight | ENSG00000101019 | UQCC1 | 20 | rs4911179 | 0.626121 | 2.15E-36 | 9.61E-13 | 5.33E-10 |

LDL Low Density Lipoproteins, TRIG Triglycerides, HDL High Density Lipoproteins, HB glycated hemoglobin, HC Hip Circumference, Chr Chromosome number, Allele Freq Allele Frequency, $P_{eQTL}$ p-value of the top associated cis-eQTL of the probe, $P_{GWAS}$ GWAS p-value of the top cis-eQTL, $P_{SMR}$ p-value for gene-trait association from the SMR test, $P_{HEIDI}$ p-value from HEIDI test to indicate whether the gene-trait association is due to a single shared genetic variant (the smaller $P_{HEIDI}$ the more likely that there are more than one genetic variant)

Conforming, in part, what was seen from the MR network, the only probes which passed the significant SMR threshold (P_SMR_ < 5.0 × 10^−6^) for the cognitive traits, were detected for the Reaction Test (processing speed domain) (Table 3, Table S4).

**Table 3:** SMR probes for the cognitive traits in Europeans which passed the SMR threshold.

| Trait | Probe ID | Gene | Chr | Top SNP | Allele Freq | $P_{\text{eQTL}}$ | $P_{\text{GWAS}}$ | $P_{\text{SMR}}$ |
| --- | --- | --- | --- | --- | --- | --- | --- | --- |
| RT | ENSG00000260075 | NSFP1 | 17 | rs35732828 | 0.160345 | 6.58E-62 | 5.58E-09 | 3.80E-08 |
| RT | ENSG00000176681 | LRRC37A | 17 | rs35732828 | 0.160345 | 2.60E-29 | 5.58E-09 | 2.29E-07 |
| RT | ENSG00000073969 | NSF | 17 | rs35732828 | 0.160345 | 2.28E-25 | 5.58E-09 | 3.67E-07 |
| RT | ENSG00000228696 | ARL17B | 17 | rs35732828 | 0.160345 | 1.96E-21 | 5.58E-09 | 6.72E-07 |

RT Reaction Test, Chr Chromosome number, Allele Freq Allele Frequency, $P_{\text{eQTL}}$ p-value of the top associated cis-eQTL of the probe, $P_{\text{GWAS}}$ GWAS p-value of the top cis-eQTL, $P_{\text{SMR}}$ p-value for gene-trait association from the SMR test, $P_{\text{HEIDI}}$ p-value from HEIDI test to indicate whether the gene-trait association is due to a single shared genetic variant (the smaller $P_{\text{HEIDI}}$ the more likely that there are more than one genetic variant)

Functional analysis of the 86 genes shows that the most of the significant hits in GO enrichment analysis are involved in MHC protein complex assembly, peptide assembly, antigen binding, Endoplasmic Reticulum (ER) biology (Figure 3A). KEGG enrichment analysis showed significant association with allograft rejection, Type II diabetes, Graft-versus-host disease, autoimmune thyroid disease (Figure 3A). Reactome pathway analysis showed immunological synapse as top hit (Figure 3A). Wikipathways analysis showed that top hit was Ebola virus infection in host (Figure 3A). Overall, these functional results mainly point towards a prominent role of the immune system in the metabolic dysfunction and neurological diseases (Figure 3A).

**Figure 3.**
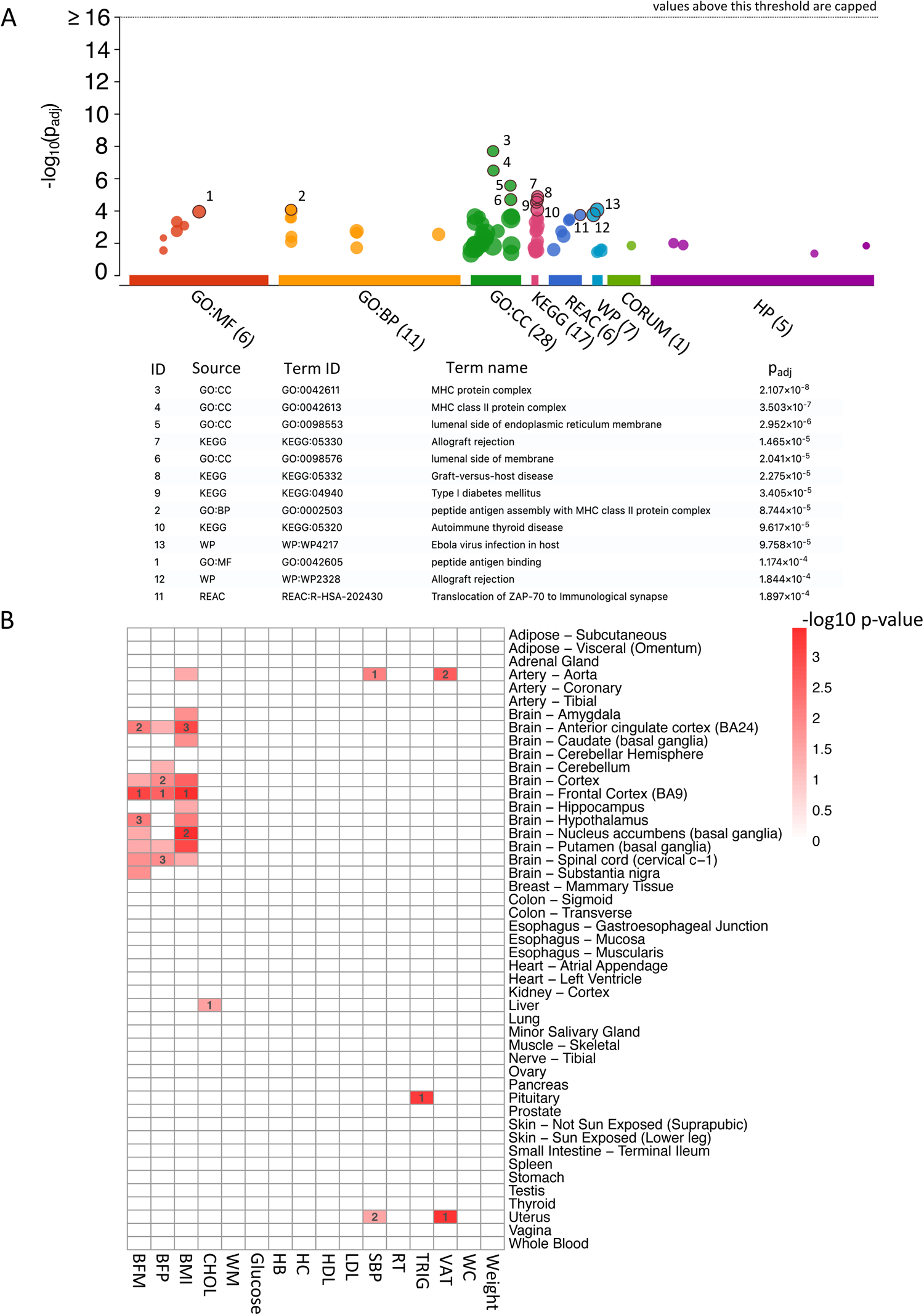
**(A)** Graphical representation of the gProfiler Gene Ontology (GO) analysis of the 86 unique significant genes detected in the SMR analysis which passed the SMR threshold (P_SMR_ < 5.0 × 10^−6^) for the European individuals. Top significant results based on adjusted p-values for Gene Ontology (GO) and pathways enrichment were selected and highlighted in black borders. GO molecular function (GO:MF); GO biological process (GO:BP); GO cellular component (GO:CC); Kyoto encyclopedia of genes and genomes (KEGG); Reactome (REAC); WikiPathways (WP); CORUM Protein Complexes (CORUM); Human Phenotype Ontology (HP). **(B)** Tissue Enrichment analysis of the GWAS summary statistics for the Europeans. Traits are on the x-axis and tissue types on the y-axis. Dark red indicate high enrichment. Numbers inside the red boxes indicate top 3 most associated tissues. Gene expression data in various tissues based on Genotype-Tissue Expression (GTEx) database. Systolic Blood Pressure (SBP), Low Density Lipoproteins (LDL), High Density Lipoproteins (HDL), Waist Circumference (WC), Hip Circumference (HC), Cholesterol (CHOL), Triglycerides (TRIG), Body Mass Index (BMI), Glycated Hemoglobin (HB), Body Fat Mass (BFM), Body Fat Percentage (BFP), Working Memory (WM), Reaction Test (RT), Visual Attention Test (VAT).

### Identification of tissue and cell types underlying susceptibility to obesity and cognitive functions

Previous reports have detected an exclusive enrichment of BMI GWAS variants in brain tissue^47^. In order to replicate this and to extend it to other metabolic and cognitive traits, we performed tissue enrichment analysis of the GWAS summary statistics with a slightly relaxed threshold (p-value < 5e-05). We observed strong association of BMI in brain tissues, especially in the frontal cortex, in addition to Body Fat Mass and Percentage (Figure 3B).

Cell-type-specific enrichment analysis (CSEA)^48^ of the BMI GWAS summary statistics in the Europeans, showed strong association with inhibitory and limbic system neurons (Figure 4A). In addition, we performed a cell type enrichment analysis^48^ of the 86 genes which passed the SMR threshold in the Europeans. We identified macrophages, oligodendrocytes and mucus secreting cells as top 3 cell types based on combined p-values (Figure 4B). For the 62 probes which passed the HEIDI cut-off, we identified gastric chief cells, fasciculata cells and oligodendrocytes as top 3 hits based on combine p-values (Figure 4C).

**Figure 4.**
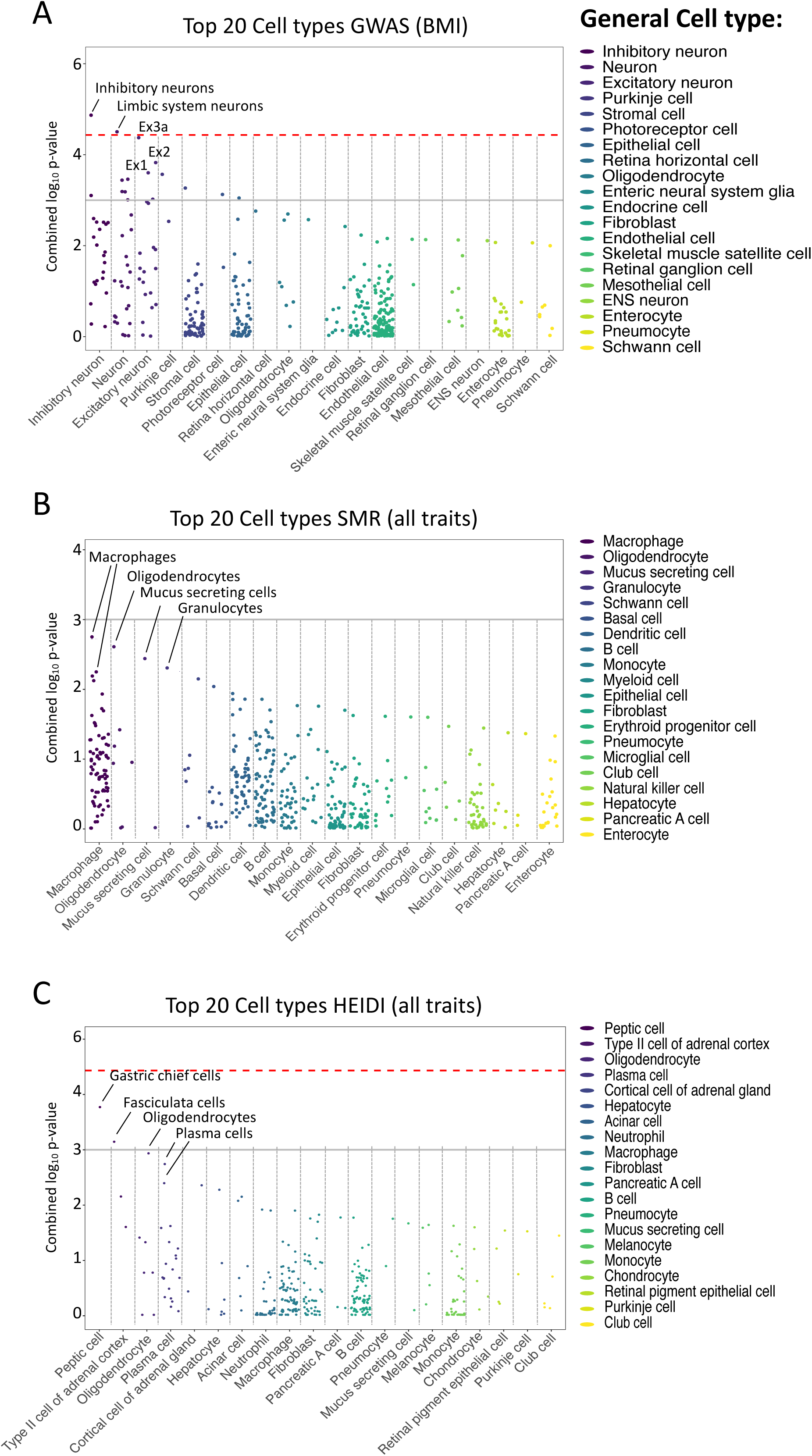
**(A)** Jitter plot of the cell-type-specific enrichment analysis (CSEA) for the top significant genes detected in the GWAS analysis for the Body Mass Index (BMI) in Europeans. **(B)** Jitter plot of the cell-type-specific enrichment analysis (CSEA) for the 86 SMR significant genes in the Europeans. **(C)** Jitter plot of the cell-type-specific enrichment analysis (CSEA) for 39 HEIDI significant genes in Europeans. Red dashed line indicates Bonferroni-corrected significance threshold (P = 3.69 × 10–5). The grey solid line indicates the nominal significance (p = 1 × 10^-3). Each dot represents one tissue-cell type in the group on the x-axis differentiated by colours. Results based on combined p-values. Top 5 general cell types are highlighted.

## Discussion

In this study, we carried out a multi-omics and genetic approach to investigate the causality between metabolic-related risk factors potentially linked to the development of dementia later in life (Figure 1). In particular, we used a network based Mendelian randomization analysis^38^ to estimate the bidirectional effects of metabolic and cognitive health in Europeans (Figure 1B). In line with previous studies^30,31^, we did not find evidence of causal association between certain glycaemic traits (e.g. glucose, glycated haemoglobin) with cognition (Figure 1B). Instead, we detected direct causal associations with traits which are more related to adiposity and lipid metabolism such as HDL and fat mass, and body shape traits such weight/waist circumference (Figure 1B). This is in agreement with a recent MR study assessing causality between abdominal adiposity and cognition in different human populations^49^. These results point towards a direct causal role of adiposity on cognitive decline as previously suggested^27,50^. In addition, in our MR analysis, we detected metabolic traits negatively influencing cognitive health and cognitive traits negatively influencing obesity (Figure 1B, Table S2). Indeed, there is evidence that metabolic-related cognitive decline is likely not just a consequence of the downstream effects of excess adipose tissue and insulin resistance, but a cause as well^50^. This results in a “vicious cycle” where lower cognitive abilities perpetuate metabolic dysfunction, which in turn perpetuate cognitive decline through the lifespan^50,51^.

As previously suggested by other reports, our tissue enrichment analysis of the top GWAS hits (Figure 3B) confirms that susceptibility to metabolic dysfunction is distributed across numerous brain areas that receive signals emanating from internal or external stimuli acting in concert to regulate feeding behaviour and energy metabolism^47,52^. In particular, these results confirm that susceptibility genes for common metabolic dysfunction may have an effect on eating addiction and reward behaviours through their high expression regions of the brains related to addition and reward such as the frontal cortex and the anterior cingulate cortex, in contrast to different pattern from monogenic obesity genes that act in the hypothalamus and cause hyperphagia^52^.

In addition, our cell type analysis of GWAS and SMR-HEIDI hits (Figure 4A-C) to investigate the identity of cell types that drive susceptibility to human metabolic dysfunction has shown strong association with different types of neurons as compared to other cell types. These results further suggest that this disfunction is distributed across multiple, mainly neuronal, cell types across the brain, thus acting on a more broadly distributed set of neuronal circuits across the brain.

We integrated the GWAS summary statistics with eQTLs data of skeletal muscle tissue by SMR and identified many reported and novel functionally relevant genes in metabolic functions and neuronal development in Europeans (Tables S3-S5).

Regarding the top hits in our SMR analysis of the Europeans, the top most significant hit was detected on the CELSR2 gene (Supp. Fig. S3A, Table S4). This gene encodes a cadherin protein with a main function in neuronal development^53^, but recently found to have a role in lipid homeostasis as well^54^. Furthermore, it has been found associated with cardiovascular diseases in the gene cluster CELSR2–PSRC1–SORT1^55^. Indeed, we detected significant signal on PSCR1 as well (Supp. Fig. S3A, Table S4). Instead, our second top hit was detected on the SNX17 gene (Supp. Fig. S3B, Table S4), recently found associated with cardiovascular diseases^56^. The gene belongs to the sorting nexin (SNX) family which consists of a diverse group of cytoplasmic and membrane-associated proteins orchestrating the process of cargo sorting^57^. Interestingly, it has been previously reported that SNX17 can regulate the trafficking of the ApoER2 receptor, member of the LDL-R family, which participates in neuronal migration during development^58^.

The majority of the significant SMR probes (33/144) (Table S4) were mapped to chromosome 16 (Figure 2A-B, Table S2), known to be associated with both obesity and neurodevelopmental disorders^43,46,59^. In particular, rearrangements on chromosome 16 including a proximal ∼200-kb deletion and a distal ∼600-kb deletion are seen in patients with the 16p11.2 deletion syndrome^60,61^. People with this disorder have developmental delay and intellectual disability and they are also at increased risk of obesity compared with the general population^45,46,61^. For example, the distal deletion carries the SH2B1 gene, a mediator of energy homeostasis, which is involved in leptin and insulin signalling^43,59^. We detected SMR signals with high significance and with suggested significance on several genes included in these deleted regions such as LAT, INO80E, FAM57B, TBX6, YPEL3, MAPK3, RP11-22P6.3, RP11-196G11.2 and BOLA2 (Figure 2A, Tables S3-S4). For example, confirming a connection between the immune system and metabolic function, the LAT (linker for activation of T cells) adaptor molecule participates in AKT activation and plays an important role in the regulation of lymphocyte maturation, activation and differentiation^62^. Instead, FAM57B is a gene which regulates adipogenesis by modulating ceramide synthesis and its haploinsufficiency contributes to changes in neuronal activity in the brain through its crucial role in lipid metabolism^63^.

We also detected significant signals on reported and novel genes on the regions flanking those deletions. For example, a well-studied gene is the Lysine Acetyltransferase KAT8 (Figure 2A). KAT8 is an important gene reported to be involved in both metabolism, being a critical regulator of central carbon metabolism, and neuron development as well^64,65^. Some of the novel significant associations are the HSD3B7 gene, which encodes an enzyme involved in the initial stages of the synthesis of bile acids from cholesterol, the Warfarin target gene VKORCl, involved in vitamin K metabolism and the two sulfotransferase SULT1A1 and SULT1A2, involved in resveratrol metabolism in adipocytes (Fig 2A). We are also detected signals on 4 long non coding RNAs (lncRNA), RP11-1348G14.4, RP11-22P6.3, RP11-196G11.2, RP11-347C12.2 (Figure 2A-B) supporting recent evidence recognizing them as contributing intermediates in obesity and inflammation^66^.

In addition, the top most significant SMR signal on chromosome 16 was detected on the well-known Fat Mass and Obesity-Associated Protein (FTO) gene which has been linked to obesity and altered connectivity of the dopaminergic neurocircuitry, being highly expressed in the hypothalamus^67^ (Figure 2B, Table S4). For this gene, we detected a specific association with BMI confirming what previously reported^47^. Previous studies reported that FTO variants are influencing gene expression levels of downstream genes in the brain such as IRX3^68,69^, a gene that plays a role in an early step of neural development, rather than FTO gene expression levels. In our analysis, we detected a very strong co-localization of GWAS signal with gene expression in the skeletal muscle (Figure 2B), thus pointing to a different mechanism of FTO gene regulation between skeletal muscle tissue and the brain. The second top most significant SMR signal on chromosome 16 was detected on the PHLPP2 gene, a phosphatase shown to be increased during obesity leading to lipid accumulation and glucose dysfunction^70^. In line with this, we detected a specific gene-trait association with LDL and cholesterol (Table S4).

Similarly to chromosome 16, microdeletions on chromosome 11 have been associated with obesity and developmental delay as well^71^. Indeed, we detected many top hits on chromosome 11 as well (Table 2, Table S4). For example, in line with a previous study showing association of the ST3GAL4 gene with plasma concentration levels of triglycerides and LDL in Europeans^72^, we detected highly significant SMR association of this gene with LDL (Supp. Fig. S3D, Table S4). Whereas, confirming the evidence that altered activities in polyunsaturated fatty acids (PUFAs) metabolism are seen in obesity and a number of chronic diseases^73^, we detected highly significant SMR signals on the FADS1 and FADS2 genes (Supp. Fig. S3C, Table S4), encoding key enzymes in PUFA metabolism.

Regarding the cognitive traits, we detected significant gene-trait associations with the Reaction Test (Table S5) on a gene cluster on chromosome 17 which includes the N-ethylmaleimide-sensitive factor (NSF) gene, which is highly expressed in the amygdala and it has been suggested to be linked with molecular mechanisms related with memory formation^74^ and two genes (ARL17B, LRRC37A) involved in the pathogenesis of the Koolen-De Vries Syndrome^75^, a developmental delay disorder in children characterised by intellectual disability. Interestingly, the Koolen-De Vries Syndrome is characterised by 17q21.31 microdeletions, the same region where we detected high SMR signal, and by truncating variants of the KAT8 regulatory NSL complex unit 1 (KANSL1) gene, which falls as well in the deleted region. KANSL1 is known to be part of the non-specific lethal (NSL) complex, which regulates global transcription by histone modification^76^. The complex includes several proteins including KAT8 on chromosome 16, on which we also detected high SMR signal as aforementioned (Figure 2A). This points to an important role of epigenetics integrity in both adipogenesis and neuronal development as recently reported^77,78^.

Overall, our functional analysis of the SMR results (Figure 3A) is in agreement with the fact that metabolic function is associated with increased MHC class II antigen presentation in adipocytes, which leads to increased proinflammatory T-cell activity in adipose tissue^79^. This generalized inflammation in adipocyte characterized by alteration in lipid metabolism would result in a dysregulation of glucose metabolism which ultimately may impact as well cognitive functions^80^.

Finally, our cell-type specific enrichment analysis (CSEA) of the SMR results, supports the existence of a “gut–brain axis” which includes interactions through the nervous system, and mutual crosstalk with the immune and the endocrine systems as previously reported^81,82^.

## Conclusions

In conclusion, this study provided novel insights into the genetic architecture of metabolic dysfunction and highlighted its putative causal relationship with cognitive decline. Results in this study can contribute to developing biomarkers, identifying drug targets and may have significant implications for global public health policies providing dietary recommendations for the management of obesity which may lead of lowering the risk of developing dementia during ageing.

## Materials and Methods

### Study Cohorts

Established with the aim of discovering genetic and non-genetic contributors to human diseases, the UK Biobank is a very large prospective cohort initiated in 2006 with an ongoing follow-up, including more than 500.000 participants living in the UK^83,84^. The cohort comprises people aged between 40-69 years with a 0.84 male/female ratio^83^. The majority of participants (94.6%) are of white British ethnicity, whereas the rest are South Asians and Black individuals^83^. Recruitment of the participants was based on proximity to assessment centres. Participants gave informed consent and provided detailed demographic, socioeconomic, lifestyle and health-related data via a touchscreen questionnaire, including medication history. As previously described, participants then underwent a large range of physical and biological assessment measures, including repeated blood pressure measurements, height and weight, from which BMI was derived, accelerometery and neuroimaging visits^83,85^; in addition to urine, saliva and blood collection for biomarker and genetic assessments as previous^84,86^.

Genotyping has been performed using the Affymetrix UK BiLEVE Axiom array on an initial 50,000 participants and the remaining 450,000 participants have been genotyped using the Affymetrix UK Biobank Axiom array^83^. Details of the quality control measures, imputation and reference genome utilised were described as previously^83^.

Furthermore, cognitive tests have been administered at several time points to UK Biobank participants. These tests were developed specifically for UK Biobank, to enable computerized administration at scale without staff involvement, and are thus non-standardized. As previously described^85^, a battery of several baseline and follow-up cognitive tests were administered in English language to participants; covering four major cognitive domains (episodic and working memory, executive function, processing speed and attention).

### Genetic Data Pre-processing

In order to run the MR analysis of the individuals with European ethnicity, we obtained genotype calls of more than 400.000 individuals, available following UK Biobank 2022 release, after centrally performing quality control (QC) procedures and imputation^83,87^. From these individuals, in order to avoid population stratification, we excluded participants with non-European ancestry. We removed from the analysis individuals for which a genetic kinship to other participants of at least one relative was identified, individuals with a missingness > 10% and with an heterozygosity ±3 SD from the mean. In addition, we removed individuals for which gender information was ambiguous or incorrect. After applying these filters, genotype calls of 261.089 individuals remained. The genotype calls consisted of 140.180 females and 120.909 males. Further stringent QC filters were applied using PLINK v.1.9^88^. Markers with a call rate < 95% (--geno 0.05), or those deviating from Hardy-Weinberg equilibrium (Bonferroni-corrected p-value threshold = 0.001) were removed. We filtered out rare variants (--maf 0.01) and we set a minor allele count (--mac) cut-off equal to 3.

### Genome-wide association study (GWAS) analysis

We estimated genome wide associations (GWAS) for several quantitative or binary metabolic and cognitive phenotypes obtained from the UK Biobank (Table S1), by fitting mixed-effects regression models accounting for population stratification and relatedness using REGENIE v2.0^89^. After QC procedures, the Genotype calls of the female dataset included 587.951 SNPs, the male dataset included 591.769 SNPs and the one altogether 577.742 SNPs. To run the GWAS analysis of the Genotype calls, we included as covariates: sex (when required), age, age^2^ and 10 ancestry-informative principal components (PCs) derived from the analysis of LD-pruned (200 variant windows, 100 variant sliding window and r^2^ < 0.2) common variants (--maf 0.01) from Genotype calls, as well 10 principal components derived from the analysis of LD-pruned (200 variant windows, 100 variant sliding window and r^2^ < 0.2) rare variants from the Genotype calls with a frequency in between a minor allele count of 3 (--mac 3) and 1% (--max-maf 0.01), in order to further avoid population stratification. We obtained PCA values by PLINK v2.0^88^. To run REGENIE, we used the female, male or altogether as input in step 1, in which the whole genome regression model is fit to the traits, and a set of genomic predictions are produced as output. The predictions are then used in step 2 where the association model was performed by including the aforementioned covariates. We run GWAS analysis on females, males or altogether.

### Mendelian Randomization (MR) Network analysis

We built the MR networks by the R package “bimmer” as previously described^38^. Briefly, to run “bimmer”, we used the male GWAS summary statistics for SNP selection and weight estimation, whereas the female GWAS summary statistics were used for model fitting. Initially, we performed pre-processing steps where we clumped the male GWAS summary statistics to *p* = 0.05 with *r*2 *<* 0.05 and distance of 500 kilobases by PLINK 1.9^88^. The clumped SNPs were then utilized to select SNPs in the female GWAS summary statistics by using a “p_thresh” value of 0.05. We used the “egger_w” method to calculate a total causal effect (TCE) matrix on the female summary statistics by utilising the selected SNPs from the male dataset. Subsequently, we filtered the TCE matrix to remove problematic phenotypes with the parameters max_SE = 0.9 and max_nan_perc = 0.9. Then, we converted the TCE matrix to a direct causal effect (DCE) matrix. To do this, we constructed at first weights with a max_min_ratio equal to 1000. Finally, we used the fit_inspire() function to fit the DCE matrix and then we obtained the estimate of the DCEs at the selected index which was used to build the network by ggnet2^38^.

### Pleiotropy analysis

We conducted an SMR&HEIDI analysis with the SMR software as previously described. (https://yanglab.westlake.edu.cn/software/smr/)^33^. To run this analysis, we utilised GWAS summary statistics obtained previously for the European individuals as outcome. We performed SMR analysis on the sex-combined for participants of European ancestry, using eQTL data as exposure. We downloaded the lite version of V8 release (n = 73-670) eQTLs summary statistics (only SNPs with P < 1e-5 were included) obtained from the GTEx Consortium^41^ for European individuals. We adopted the default settings in SMR (e.g., peqtl-smr = 5.0e-8, peqtl-heidi = 1.57e-3, diff-freq = 0.2, diff-freq-prop = 0.05, cis-window = 2000 kb, minor allele frequency [MAF] > 0.01, removing SNPs in very strong linkage disequilibrium [LD, r2 > 0.9] with the top associated eQTL, and removing SNPs in low LD or not in LD [r2 less than 0.05] with the top associated eQTL).

We also conducted the heterogeneity in dependent instruments (HEIDI) test to evaluate the existence of linkage in the observed association^33^.

We performed functional enrichment analysis with the R packages gprofiler2^90^ and clusterProfiler^91^ with Bonferroni correction. We built a PPI network with the online database STRING v11.5 (https://string-db.org/)^92^.

### Integration with RNA-seq Public Data

We performed a tissue enrichment analysis of the sex-combined GWAS summary statistics of the metabolic and cognitive traits, with the deTS R package^93^. To run the enrichment analysis, phenotypes with less than 20 genes were excluded and we utilised bulk RNA-seq gene expression data from the GTEx database^41^.

We performed a cell-type-specific enrichment analysis (CSEA) of the sex-combined GWAS summary statistics of the metabolic and cognitive traits with the online web-based application WebCSEA^48^, which contains a total of 111 single cell RNA-seq (scRNA-seq) panels of human tissues across 11 human organ systems.

## Data Availability

The UKBB phenotypic data and genetic data analysed during the current study are available from UK Biobank. GWAS pre-processing and analysis was performed on the UK Biobank Research Analysis Platform (RAP) following UK Biobank coding guidelines (https://dnanexus.gitbook.io). Bimmer is implemented as an open-source R package available at https://github.com/brielin/bimmer. The deTS tissue enrichment tool is implemented as an open-source R package available at https://github.com/bsml320/deTS.

## Online resources

UK Biobank https://www.ukbiobank.ac.uk/

https://dnanexus.gitbook.io

SMR https://yanglab.westlake.edu.cn/software/smr/

g:Profiler https://biit.cs.ut.ee/gprofiler/gost

WebCSEA https://bioinfo.uth.edu/webcsea/

STRING https://string-db.org/

## Funding Statement

The research has been conducted using the UK Biobank Resource (project # 43769). UK Biobank was established by the Wellcome Trust medical charity, Medical Research Council, Department of Health, Scottish Government and Northwest Regional Development Agency. It also had funding from the Welsh Assembly Government, British Heart Foundation and Diabetes UK. The authors are grateful to the UK Biobank participants for making such research possible. This work was supported by the Singapore Ministry of Education under its Singapore Ministry of Education Academic Research Fund Tier 1.

## Author Contributions

P.A.L designed methodology, performed the analyses and summarized the results, writing code and data preparation, writing-original manuscript draft. S.R.L conceptualized the study, funding acquisition, designed methodology, resources acquisition, supervised the study and reviewed and edited the manuscript.

## Conflicts of Interest

All authors declare no conflict of interest.

## Acknowledgments

This research has been conducted using the UK Biobank Resource under Application Number # 43769 and uses data provided by patients and collected by the NHS as part of their care and support. We thank the UK Biobank (UKBB) for their assistance in preparing the data and providing access to computational resources. Part of the analyses were conducted on the Research Analysis Platform (http://ukbiobank.dnanexus.com). We are grateful to the National Supercomputing Center, Singapore (https://www.nscc.sg), where part of this work was performed. We thank Matthew Tham and the members of the Integrative Biology of Disease Group for their insightful comments and suggestions.

---

**Fig S1.**
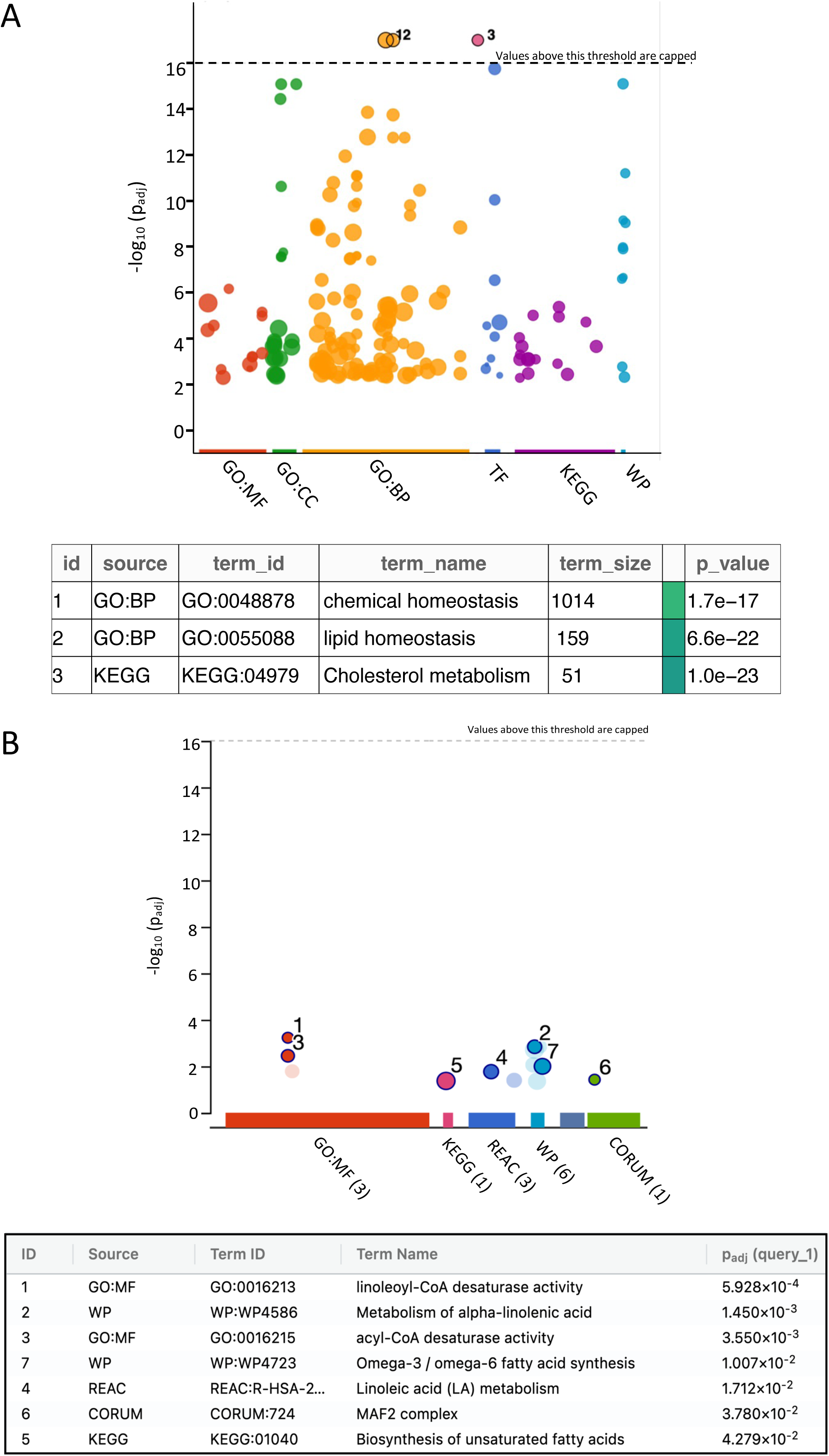
**(A)** Plot of the gProfiler ontology analysis of the significant GWAS hits for both the metabolic and cognitive traits which passed the threshold (p-value < 5e-08) for the European individuals. **(B)** Plot of the gProfiler ontology analysis of the 39 unique significant genes that passed both the SMR and HEIDI test in the SMR analysis for the European individuals. GO molecular function (GO:MF); Kyoto encyclopedia of genes and genomes (KEGG); Reactome (REAC); WikiPathways (WP); CORUM Protein Complexes (CORUM).

**Fig S2.**
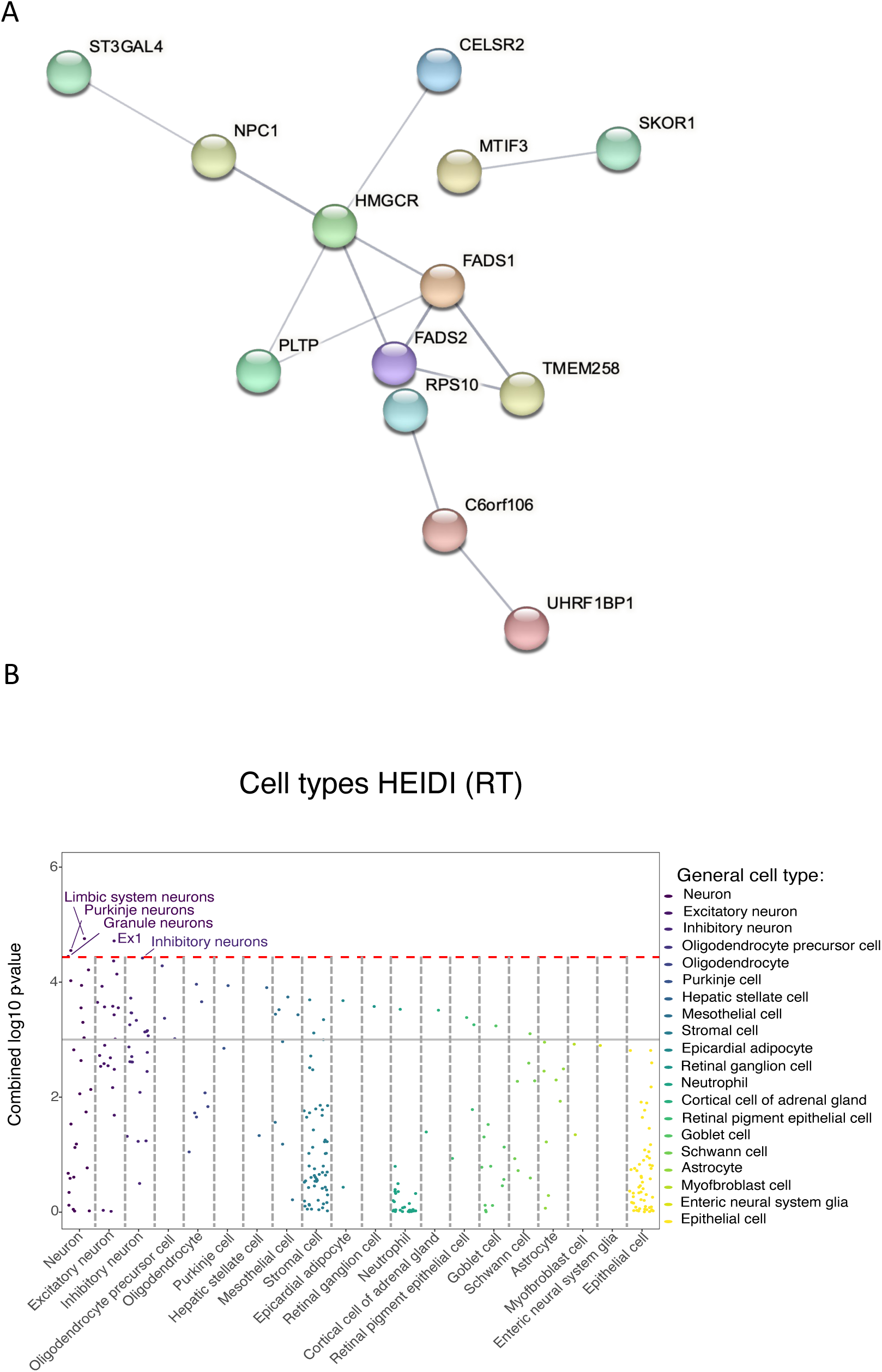
**(A)** Protein-Protein Interaction (PPI) network generated by STRING database of the 39 unique significant genes which passed both the SMR and HEIDI threshold in Europeans. **(B)** Jitter plot of the cell-type-specific enrichment analysis (CSEA) for the GWAS significant genes detected in the Reaction Test (RT) trait of the Europeans individuals. Red dashed line indicates Bonferroni-corrected significance threshold (P = 3.69 × 10–5) by 1355 TCs. The grey solid line indicates the nominal significance (p = 1 × 10^-3). Each dot represents one tissue-cell type in the group on the x-axis differentiated by color. Results based on combined p-values.

**Fig S3.**
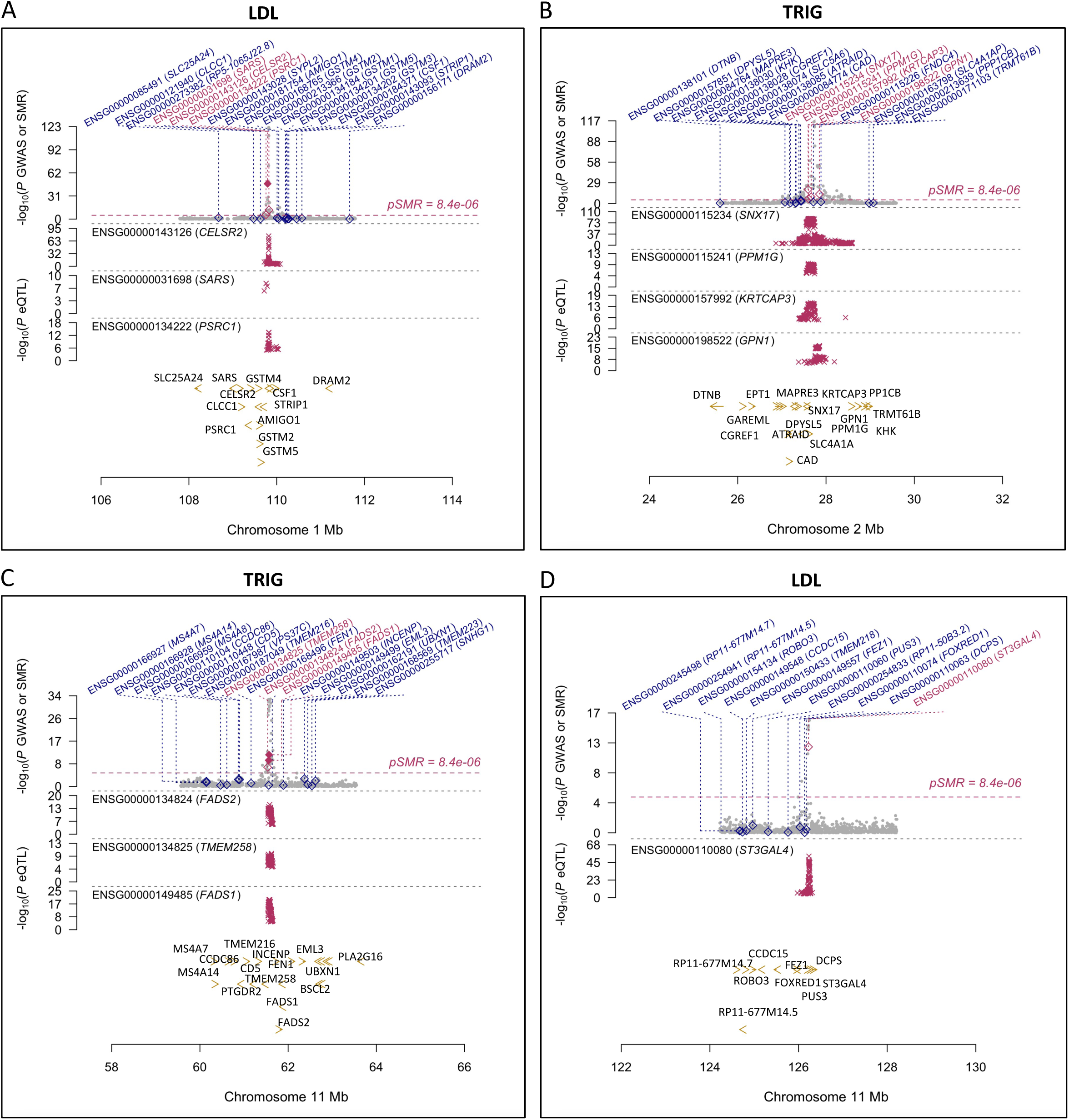
**(A)** SMR locus plot of the CELSR2 locus for the Low-Density Lipoproteins (LDL) trait in Europeans. **(B)** SMR locus plot of the SNX17 locus for the Triglycerides (TRIG) trait in Europeans. **(C)** SMR locus plot of the FADS2 locus for the Triglycerides (TRIG) trait in Europeans. **(D)** SMR locus plot of the ST3GAL4 locus for the Low-Density Lipoproteins (LDL) trait in Europeans. On the y-axis, P-values for SNPs reported in this GWAS meta-analysis are represented by grey dots and diamonds represent the P-values for probes from the reverse SMR test. Magenta crosses in the middle plot represent the cis-eQTL P-values of SNPs associated with gene expression in skeletal muscle (GTEx). Bottom yellow arrows indicate genes over the genome positions in hg38 (x-axis).

## Notes

### Competing Interest Statement

The authors have declared no competing interest.

### Author Declarations

The Research Ethics Committee of the UK Biobank gave ethical approval for this work through the approval of application project # 43769.

