## Supplemental Table 1 for "Multi-omics and Mendelian Randomization network analysis of the association between metabolic and cognitive functions in the UK Biobank Database"

| ID UKB METABOLIC | FULL NAME | ID UKB COGNITIVE | FULL NAME |
|---|---|---|---|
| SBP | Systolic Blood Pressure | WM | Working Memory |
| DBP | Diastolic Blood Pressure | EM | Episodic Memory |
| HDL | High Density Lipoproteins | FIQ | Fluid IQ |
| LDL | Low Density Lipoproteins | RT | Reaction Test |
| Glucose | Glucose | VAT | Visual Attention Test |
| WC | Waist Circumference | | |
| HC | Hip Circumference | | |
| TRIG | Triglycerides | | |
| Weight | Weight | | |
| HB | Glycated Hemoglobin | | |
| BMI | Body Mass Index | | |
| VATV | Visceral Adipose Tissue Volume | | |
| VATM | Visceral Adipose Tissue Mass | | |
| BFP | Body Fat Percentage | | |
| BFM | Body Fat Mass | | |
| AATV | Abdominal Adipose Tissue Volume | | |
| CHOL | Cholesterol | | |
